# Functional anomaly mapping lateralizes temporal lobe epilepsy with high accuracy in individual patients

**DOI:** 10.1101/2023.02.05.23285034

**Authors:** Taha Gholipour, Andrew DeMarco, Xiaozhen You, Dario J Englot, Peter E Turkeltaub, Mohamad Z Koubeissi, William D. Gaillard, Victoria L Morgan

## Abstract

**Objective:** Mesial temporal lobe epilepsy (mTLE) is associated with variable dysfunction beyond the temporal lobe. We used functional anomaly mapping (FAM), a multivariate machine learning approach to resting state fMRI analysis to measure subcortical and cortical functional aberrations in patients with mTLE. We also examined the value of individual FAM in lateralizing the hemisphere of seizure onset in mTLE patients. Methods: Patients and controls were selected from an existing imaging and clinical database. After standard preprocessing of resting state fMRI, time-series were extracted from 400 cortical and 32 subcortical regions of interest (ROIs) defined by atlases derived from functional brain organization. Group-level aberrations were measured by contrasting right (RTLE) and left (LTLE) patient groups to controls in a support vector regression models, and tested for statistical reliability using permutation analysis. Individualized functional anomaly maps (FAMs) were generated by contrasting individual patients to the control group. Half of patients were used for training a classification model, and the other half for estimating the accuracy to lateralize mTLE based on individual FAMs. Results: Thirty-two right and 14 left mTLE patients (33 with evidence of hippocampal sclerosis on MRI) and 94 controls were included. At group levels, cortical regions affiliated with limbic and somatomotor networks were prominent in distinguishing RTLE and LTLE from controls. At individual levels, most TLE patients had high anomaly in bilateral mesial temporal and medial parietooccipital default mode regions. A linear support vector machine trained on 50% of patients could accurately lateralize mTLE in remaining patients (median AUC =1.0 [range 0.97-1.0], median accuracy = 96.87% [85.71-100Significance: Functional anomaly mapping confirms widespread aberrations in function, and accurately lateralizes mTLE from resting state fMRI. Future studies will evaluate FAM as a non-invasive localization method in larger datasets, and explore possible correlations with clinical characteristics and disease course.

## Introduction

The pathophysiology of mesial temporal lobe epilepsy (mTLE) involves neuronal dysfunction in both mesial temporal structures and distant brain regions through anatomical and functional pathways.[1-6] The extent of epileptic networks may explain individual variability in ictal semiology and interictal neurocognitive deficits in focal epilepsy.[7-10] Network alterations and properties may also guide seizure focus localization, a common challenge in presurgical evaluations for focal epilepsies.[11, 12] Whole brain fluctuations in neuronal activity can be indirectly measured by resting state functional MRI (rsfMRI).[13, 14] In the commonly used functional connectivity (FC) approach, covariance between pairs of brain regions are calculated to represent network organization. While simple and reproducible, the bivariate approach may overlook concurrent changes outside of each tested pair. Estimating dynamic changes in signal correlations is another FC limitation that is partially addressed by using a sliding time window and bivariate connectivity analysis.[15] At a group level, FC can reveal largescale local and distant network alterations in TLE,[4, 6, 12, 16-18] but efforts for individual characterization and classification of epilepsy using FC has led to modest results.[12, 19, 20] Other studies have used univariate analysis methods of rsfMRI analysis such as amplitude of low frequency fluctuation (ALFF), which measures each regional signal fluctuations independently. Group-level changes in ALFF are also detected in mesial temporal and default mode network, with different properties in right and left onset.[21, 22] Functional anomaly mapping, unlike previous bivariate and mass-univariate methods, is a multivariate analysis machine learning approach to rsfMRI and has the advantage of accounting for concurrent changes in all regions. The result is a group or individual functional anomaly map (FAM). The method has been validated for studying cerebrovascular lesions and their effects on distant brain function,[23] and showed correlation with clinical symptoms and cognitive profiles.[24] For generating an individual FAM, time-varying rsfMRI signal aberrations are estimated by contrasting individual patient data to a group of controls in a support vector regression model (Figure 1). To date, limitations in measuring individual functional variability and aberrations that correlate with clinical characteristics has hindered clinical translation and remains a crucial gap for development of biomarkers and diagnostic tools. In this study, we used FAM to map cortical and subcortical rsfMRI anomalies in mTLE at group and individual levels. We hypothesized that 1) group FAM analysis would detect distant anomalies in addition to the region around the seizure onset zone; 2) the pattern of anomaly is different in left and right mTLE at both group and individual FAM levels; and 3) this difference in individual anomaly pattern can be used to lateralize mTLE patients. We tested those hypotheses by generating group and individual FAMs for patients with mTLE, and analyzed the results in relation to relevant clinical characteristics.

**Fig. 1.**
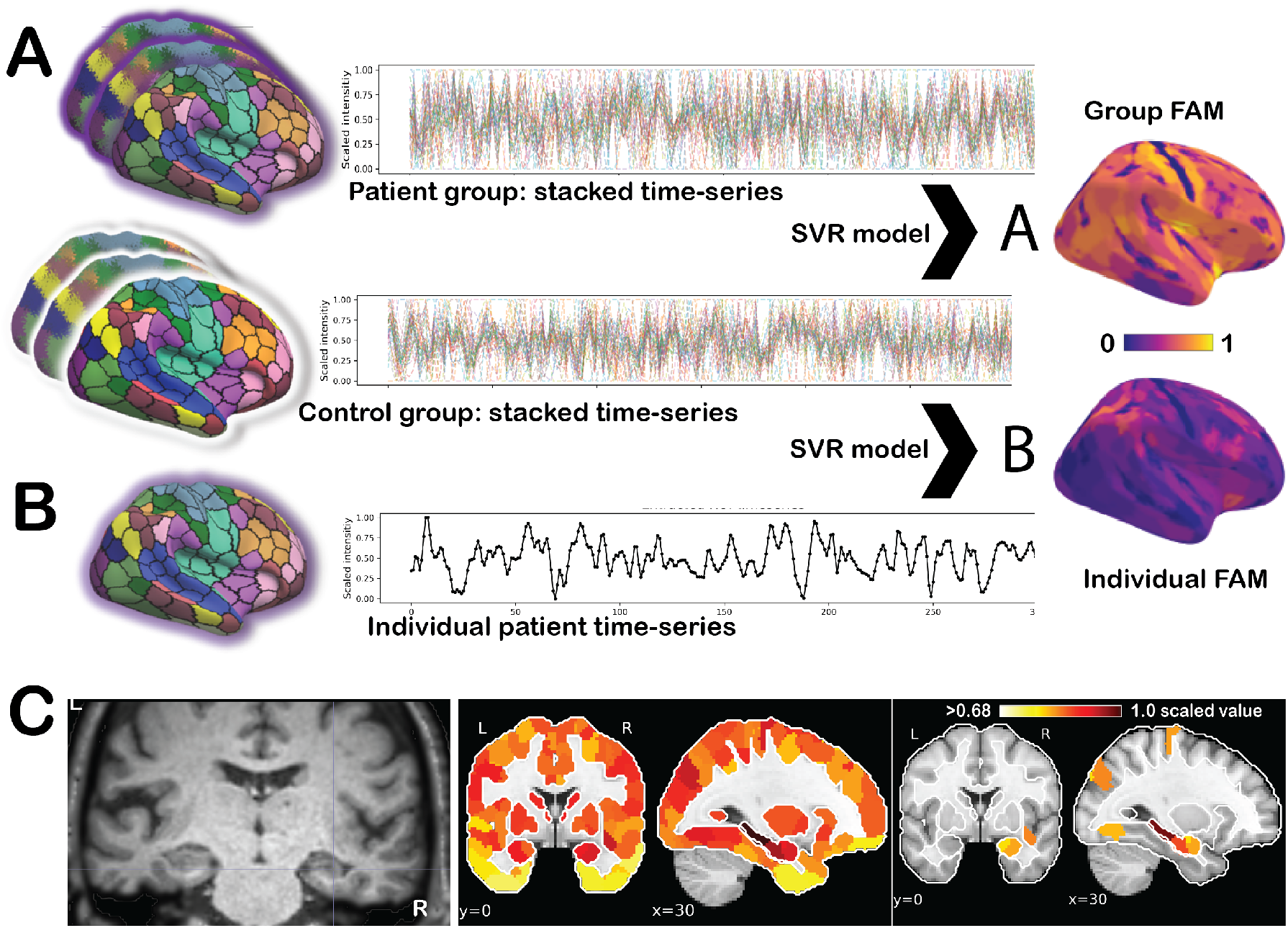
Functional anomaly mapping framework. Time-series of resting state functional MRI are extracted for each participant, scaled between 0 and 1 and flattened as a space-time vector. Stacked time-series from the control group (middle row, each line representing all ROIs for one person) are used as the normative control sample in support vector regression (SVR) models. Time-series of a group of patients (A) or single patient (B) are contrasted with the control group to generate group (A) or individual (B) analysis. Results for each ROI are averaged over the time dimension and are back-projected to same brain space for visualization. The mean of the model weights is scaled between 0 and 1, with 1 representing the ROI with the highest importance in distinguishing a patient group or a single patient from the control group. Panel (C) illustrates a coronal slice of T1-weighted image of a patient with right hippocampal sclerosis, with right hippocampal atrophy. Individual FAM of the same patient projected to MNI space is illustrated without threshold (middle image) and 0.68 (right coronal and sagittal panels). Note the anomaly in several ROIs from the sclerotic hippocampus.

## Methods

### Participants

We used a single-center dataset of adult rsfMRI from Vanderbilt University Medical Center for this study. The diagnosis and lateralization of mTLE in patients were determined by the clinical team through interictal and ictal recording, clinical epilepsy-protocol 3T MRI, fluorodeoxyglucose positron emission tomography (FDG-PET), and intracranial recording, when indicated. We included patients with and without pre-surgical MRI evidence of hippocampal sclerosis (HS) based on expert neuroradiologist review. Data from 94 adult healthy controls were used as the normative group for generating FAM.

### MRI acquisition and preprocessing

All participants were scanned using a 3T Philips Achieva MRI with a 32channel head coil. This included a 1-mm isotropic T1weighted anatomical and two consecutive runs of T2*weighted fMRI scans, with the participant instructed to rest with eyes closed. Each functional scan lasted 10 min with the following parameters: voxel size= 3×3×3.5 mm, 34 slices with a 0.5 mm slice gap, TR = 2000 ms, TE=35 ms. Anatomical and functional image preprocessing was performed using the fMRIPrep pipeline (version 20.2).[25] Functional images were co-registered with the anatomical image with nine degrees of freedom, followed by estimation of head-motion parameters, and resampling of preprocessed time-series into standard space (MNI152NLin2009cAsym, 2 mm resolution). We used xcpEngine (version 1.2.1) with 36-parameter denoising design (six motion parameters, white matter, cerebrospinal fluid, and global signal time-series, their temporal derivatives and quadratic terms) plus movement spikes as regressors, to extract time-series of regions determined by parcellation atlases, and used a band-pass filter of 0.01-0.08 Hz.[26, 27] Cortical regions of interest (ROIs) were determined by the 400-parcel Schaefer atlas.[28] Subcortical ROIs were determined by the 32-ROI Melbourne Subcortex atlas. [29] Both atlases are derived from functional human brain connectivity, hence providing network context to our analysis. The Schaefer atlas is derived from normative local and global FC measures with each parcel falling into one the seven large-scale functional networks,[28, 30] and the Melbourne Subcortex atlas delineates functional (as opposed to traditional anatomical) subsegments in TLE-relevant thalamus, hippocampus, and amygdala.

### Functional anomaly mapping framework

We developed a functional anomaly mapping framework based on the methods described and validated by DeMarco and Turkletaub.[23] Their method involves voxel-level FAM to show aberrations in rsfMRI signal of patients with cerebrovascular lesions. Here, we adapted FAM for ROI-level analysis using support vector regression (SVR), a support vector machine implementation of multiple linear regression,[31] to fit models distinguishing patient(s) from controls. For each participant, the resting state time-series are scaled and flattened to a one-dimensional space-time (ROI-time) vector. Data vectors from the control group are stacked, generating a normative matrix with each row representing a participant and each column representing space-time values. Similarly, data vectors from a single patient (for generating individual FAM) or a group of similar patients (for group FAM) are added. An SVR model is applied to functional data by contrasting patient(s) with the control group (Figure 1). The model’s solution results in a value for each space-time, representing its importance in distinguishing the patient(s) from the control group. For visualization and comparability between individual patients, the mean of the absolute FAM values for each ROI over time is scaled (0< FAM <1; with 1 representing the ROI with the highest importance), and back-projected to standard atlas space for visualization of FAM values.

### Group FAM analysis

To test whether group FAM analysis detects distant anomalies in addition to the mesial temporal region, we generated separate models for LTLE and RTLE group analysis. The cortical and subcortical ROIs were included in the same model. We performed random permutation analyses for inferring the statistical reliability and significance for each of the two groups. Group results are tested relative to what would be obtained under the null hypothesis of no difference between patients and controls.[32] This is a non-parametric approach that compares results to a null distribution produced by randomly shuffling the labels (control or TLE) and repeating the model 5,000 times. For multiple comparisons correction, we used a continuous permutation-based family-wise error rate (cFWER) correction method, with the critical threshold calculated based on the 95th percentile of the 10th most extreme permutation P-value (v = 10) observed across each permuted result map.[33] Regions with P-values less than the critical threshold determined by cFWER were reported as significant after correction for multiple comparisons.

### Individual FAM analysis

To identify the patterns of anomaly in mTLE patients, we defined regions with high anomaly as the 86 ROIs with highest relative FAM values within each individual FAM (2% of 432 ROIs). To identify the common regions showing high anomaly among patients of each TLE group, we aggregated individual FAMs to create heatmaps for LTLE and RTLE patients and tabulated the number of patients showing high anomaly in those regions by group. To examine other factors, we compared the prevalence of those regions by presence of HS and surgical outcomes, when available. Classification using individual FAM We examined whether individual FAMs predict the hemisphere of seizure onset (left vs right). Outcomes from surgical treatment of mTLE were labeled as either Engel class I (no disabling seizures 12 months after surgery) or classes II-IV (recurrence of disabling seizures after 12 months)[34]. To show if the hemisphere of seizure onset can be inferred by the pattern of individual FAM, we trained a support vector machine (SVM) as a multivariate classifier on 50% of the individual patient FAM map data with the following parameters: linear kernel, C=1.0, with 3-fold cross-validation. We estimated the model’s performance by resampling and splitting data to train and test subset 1000 times, and reported the median and range of accuracy and area under the curve (AUC) of receiver operating characteristic curve (ROC) on the testing group. The resampling for training was stratified by their classification group (for example left or right) to avoid bias in performance. We reported the regions with the highest importance in predicting laterality from the trained SVM, and visually compared the distribution of FAM values for those high-importance ROIs. Our dataset had an imbalance in Engel class I versus Engel class II-IV surgical outcomes, which is expected in treatment of TLE, however we examined whether a model similar to lateralization SVM can classify patients to surgical outcomes groups based on their individual FAM. Finally, given the high dimensionality of FAM data, to provide a more interpretable visualization of the distance and relationships between patients, we applied principal component analysis (PCA)[35] to the set of individual patient FAMs. We plotted patients based on their first two principal components, and the probability of the distribution was demarcated as kernel density estimation. If SVM results a high accuracy in classification, we expect a visual separation of patients based on their groups in the low-dimensional space.

### Standard Protocol Approvals, Registrations, and Patient Consents

The Vanderbilt University Institutional Review Board approved the use of human subjects for this study and written consent was acquired from all subjects prior to participation. Analysis of anonymized data was approved by the George Washington University Institutional Review Board as exempt.

### Data availability and method reproducibility

Anonymized data and code will be shared upon request by qualified investigators. We used MATLAB (R2022a) for generating FAM results, with SVR performed using the fitrsvm function (BoxConstraint=1; OutlierFraction=0.05; Epsilon = 0.1; and ISDA solver). We used Python (version 3.9.7) Scikit-learn and nilearn libraries[35] for subsequent analysis steps library for visualizations.

## Results

### Participants

Fourteen LTLE and 32 RTLE patients with clinically confirmed diagnosis of mTLE and adequate image quality were included. Of those, 13 LTLE and 20 RTLE patients had MRI evidence of HS, which was confirmed by post-surgical pathology in all cases but one LTLE patient who underwent laser ablation. The majority (43/46) of patients underwent selective amygdalohippocampectomy or standard anterior temporal resection on diagnosed side of mTLE, and 39 had Engel seizure outcome 12 months after the surgery. One RTLE patient underwent responsive neurostimulation placement, with 12-month Engel score of IV, and one LTLE patient had laser ablation of amygdala and hippocampus, and achieved 12-month Engel score of I (free of disabling seizures). One patient deferred intervention. Table 1 summarizes other relevant clinical and demographic characteristics.

**Table 1.** Demographic and clinical characteristics of participants.

|  | RTLE | LTLE | Control |
| --- | --- | --- | --- |
| N (Female:Male) | 32 (17:15) | 14 (4:10) | 94 (45: 49) |
| With evidence of HS on MRI(Confirmed HS on pathology) | 20 (20) | 13 (12) | – |
| Without evidence of HS on MRI(Features of HS on pathology) | 12 (4) | 1 (1) | – |
| Mean age at scan date in years (range) | 40.8 (23-62) | 36.9 (18-68) | 38.1 (18-71) |
| Median age of onset in years | 20 | 18.5 | – |
| Handedness (Right:Left) | 25:7 | 12:2 | 81:13 |
| Resective surgery*(Engel outcome at 12-month) | 30 (27) | 13 (12) | – |

### Functional anomaly mapping shows distributed aberrations in group analysis

Group FAM results showed distributed spatiotemporal functional alterations beyond the mesial temporal and the limbic network for both RTLE and LTLE groups (Figure 2).

**Fig. 2.**
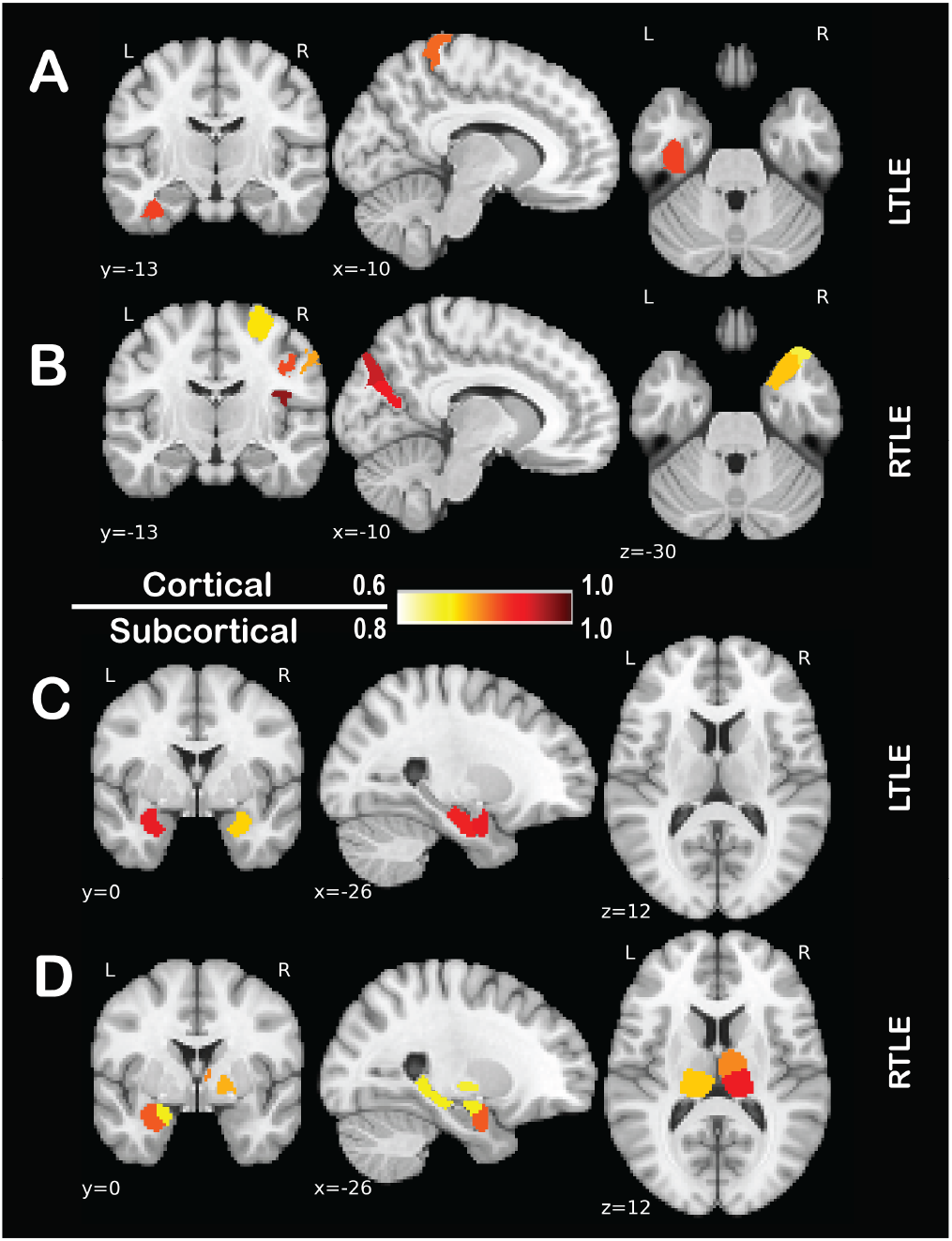
Group level anomalies in mesial temporal lobe epilepsy. In group functional anomaly mapping (FAM), temporal lobe epilepsy patients are contrasted with controls and tested for significance using permutation testing. Cortical regions (A and B) with significant group-level anomaly after accounting for multiple comparisons are illustrated (critical p-value of 0.008 for LTLE and 0.009 for RTLE). Subcortical regions had higher FAM values (C and D), however did not survive correction for multiple comparisons (shown here uncorrected p-value<0.05). Color bars are adjusted for showing different range of the scaled FAM values.

From permutation testing of group results for statistical reliability, 46 ROIs (one subcortical and 45 cortical) in RTLE, and 28 (three subcortical and 25 cortical) in LTLE showed higher anomaly compared to the control group (ROI-wise uncorrected p<0.05). Following correction for multiple comparisons, 10 cortical regions RTLE and four cortical regions in LTLE remained significant (cFWER corrected p-value threshold for V=10 of 0.009 for RTLE, and 0.008 for LTLE). Table 2 lists the ROIs with significant anomaly for each TLE group. Despite exhibiting higher FAM values relative to cortical ROIs, subcortical regions were seldom found significant using this permutation test. for example, combined hippocampus and amygdala regions had an average FAM of 0.66 in left and 0.71 in right hemisphere, while the average cortical ROIs in limbic network were 0.30 in both hemispheres.

**Table 2.**
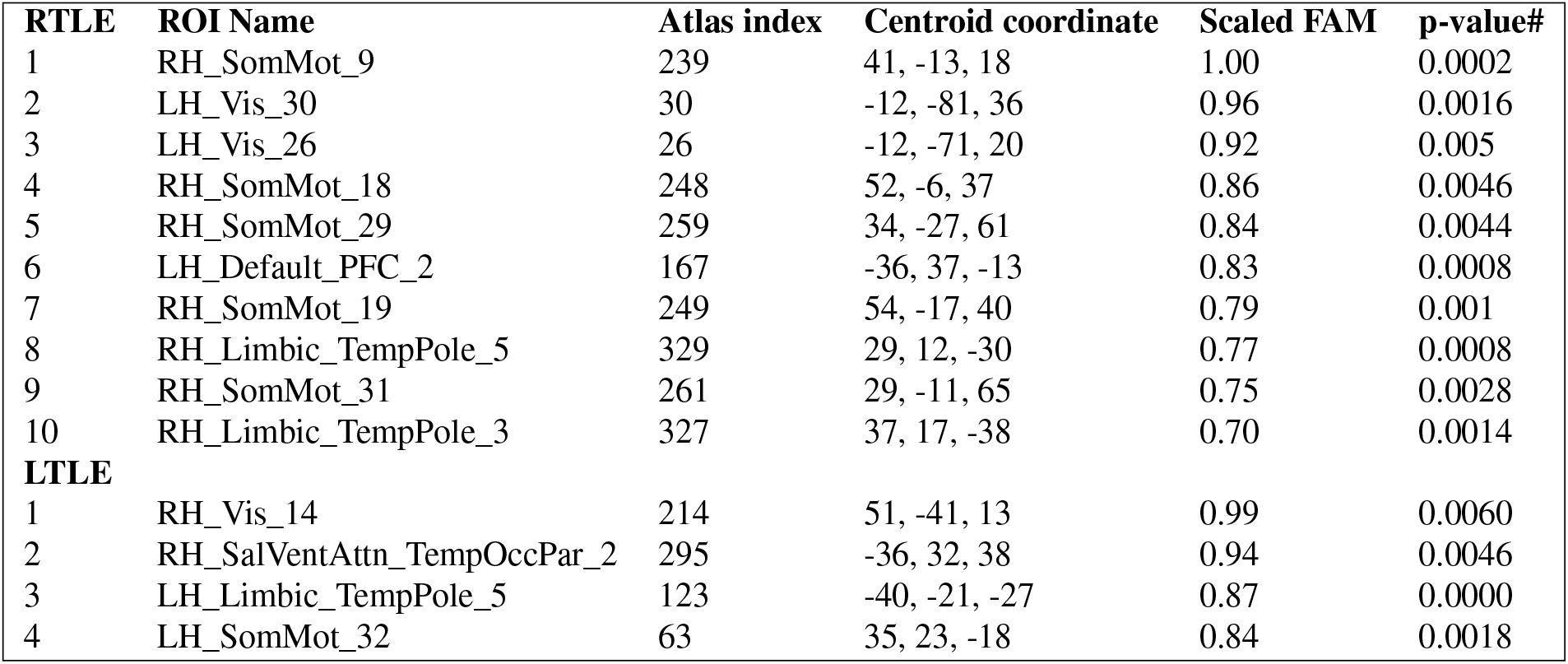
Regions of group anomaly by comparing TLE-HS groups to controls. Cortical regions with the largest measured anomaly based on group functional anomaly maps (FAM value) for right and left temporal epilepsy (RTLE and LTLE) in contrast to controls, after accounting for multiple comparisons. Region of interest (ROI) names, atlas index, and Montreal Neurological Institute centroid coordinates (R, A, S) are according to the updated atlases.[28] Each name represents hemisphere (LH: left, RH: right), network name based on Yeo 7-network affiliation of the ROI,[30] and component name. P-values are calculated based on the distribution of 5,000 permutations, with a threshold determined based on continuous family-wise error correction[33] (# p-value < 0.009 for RTLE and p < 0.008 for LTLE). None of the subcortical regions with p<0.05 met the correction threshold.

| RTLE | ROI Name | Atlas index | Centroid coordinate | Scaled FAM | p-value# |
| --- | --- | --- | --- | --- | --- |
| 1 | RH_SomMot_9 | 239 | 41, -13, 18 | 1.00 | 0.0002 |
| 2 | LH_Vis_30 | 30 | -12, -81, 36 | 0.96 | 0.0016 |
| 3 | LH_Vis_26 | 26 | -12, -71, 20 | 0.92 | 0.005 |
| 4 | RH_SomMot_18 | 248 | 52, -6, 37 | 0.86 | 0.0046 |
| 5 | RH_SomMot_29 | 259 | 34, -27, 61 | 0.84 | 0.0044 |
| 6 | LH_Default_PFC_2 | 167 | -36, 37, -13 | 0.83 | 0.0008 |
| 7 | RH_SomMot_19 | 249 | 54, -17, 40 | 0.79 | 0.001 |
| 8 | RH_Limbic_TempPole_5 | 329 | 29, 12, -30 | 0.77 | 0.0008 |
| 9 | RH_SomMot_31 | 261 | 29, -11, 65 | 0.75 | 0.0028 |
| 10 | RH_Limbic_TempPole_3 | 327 | 37, 17, -38 | 0.70 | 0.0014 |
| **LTLE** |  |  |  |  |  |
| 1 | RH_Vis_14 | 214 | 51, -41, 13 | 0.99 | 0.0060 |
| 2 | RH_SalVentAttn_TempOccPar_2 | 295 | -36, 32, 38 | 0.94 | 0.0046 |
| 3 | LH_Limbic_TempPole_5 | 123 | -40, -21, -27 | 0.87 | 0.0000 |
| 4 | LH_SomMot_32 | 63 | 35, 23, -18 | 0.84 | 0.0018 |

### Individual FAM features

We defined the high anomaly regions as each individual’s 20% highest FAM values. By counting their presence within the same patient group, bilateral mesial temporal ROIs were frequently the most anomalous regions in both LTLE and RTLE patients. In addition, most TLE patients had high anomaly in bilateral medial parietooccipital regions, most associated with visual network and posterior cingulate/precuneus anatomical regions. Figure 3 shows those regions in LTLE, RTLE, and RTLE when limited to those with HS or Engel score II-IV.

**Fig. 3.**
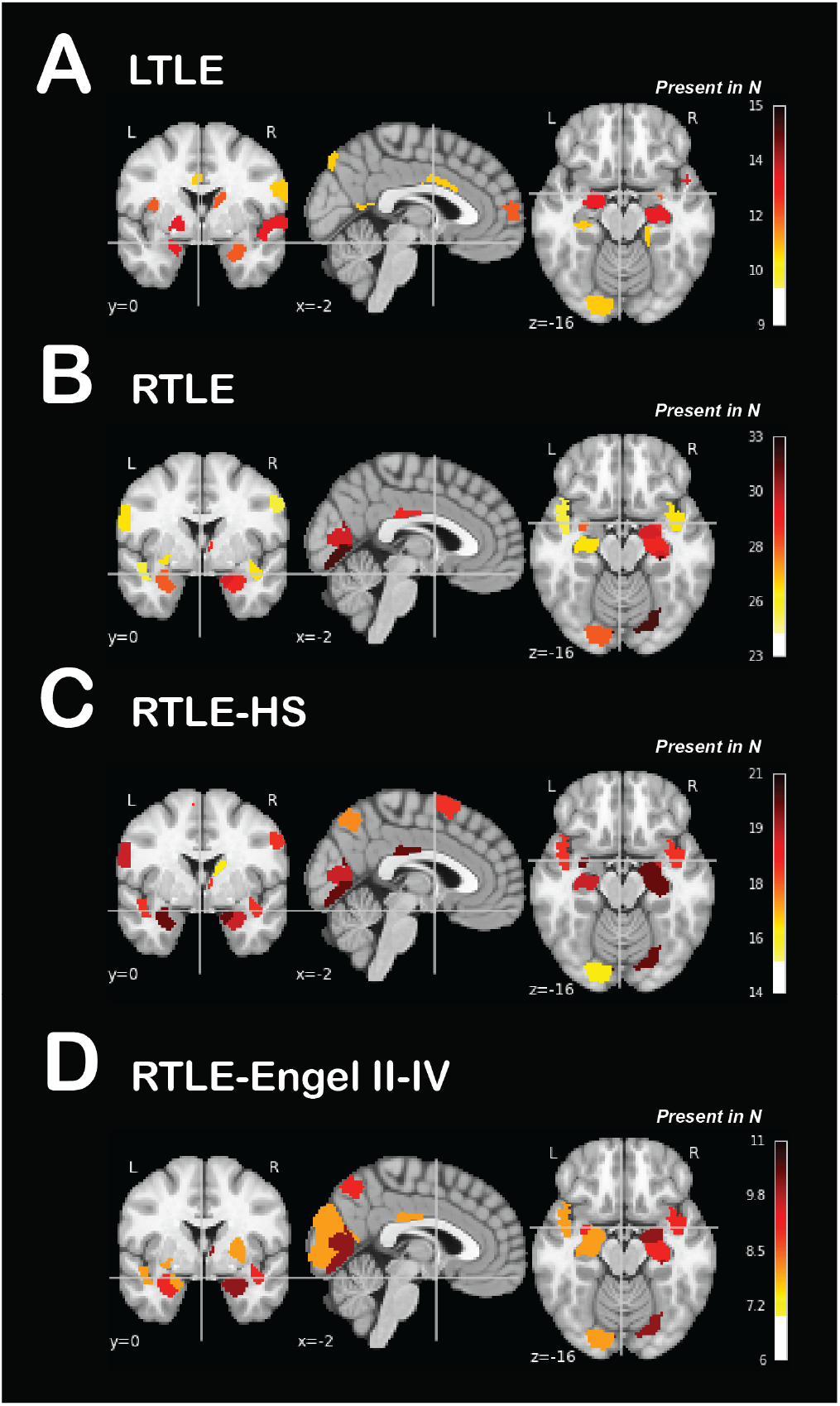
Individual anomaly heatmaps. Cumulative individual FAM heatmaps of the regions with highest anomaly (highest 20% FAM values), color map is scaled to represent proportion of patients with high anomaly in those regions: A and B) left (LTLE, N=14) and right (RTLE, N=32); C) RTLE limited to those with hippocampal sclerosis on MRI (RTLE-HS, N=20); D) RTLE subgroup with Engel score II-IV (recurrence of seizure) within 12 months from surgery (N= 10)..

### Lateralization from individual FAM patterns

The linear SVM model trained on 50% of individual FAM results predicted laterality of the other untrained 50% of data with high accuracy, with median AUC of 1.0 (inter-quartile range 0.99-1.0 across 1000 reiterations) and median accuracy of 96.87% (inter-quartile range 92.86-100%). The anomaly in the right posterior hippocampus, left temporal cortex (LH_Limbic_TempPole_5 [-40,-21,-27]), and a number of default mode network members, including medial parietooccipital regions of posterior cingulate cortex and precuneus played higher importance relative to other regions in this classification, based on their higher absolute SVM classification coefficients. Figure 4B illustrates the FAM values for selected ROIs with the highest importance. The range of individual FAM in subcortical regions did not clearly characterize the two groups or HS status. Subcortical regions showed higher anomaly, but their FAM ranges for RTLE and LTLE highly overlapped. In contrast, cortical regions showed better differentiation in range of FAM values between LTLE and RTLE patients. Visualization of the first two principal component of individual FAMs (accounting for 71% of the variance in our data) showed low overlap between the RTLE and LTLE patients and their distribution probability in the low dimensional space, irrespective of the MRI lesion status. The SVM model to classify Engel outcomes in 41 available patients (Engel class I vs. Engel class II-IV) resulted a high variance in accuracy when testing over 1000 reiterations, achieving a performance no greater than chance. A similar SVM model trained to classify patients based on their HS status did not achieve greater than chance performance in our imbalanced data (see Table 1).

**Fig. 4.**
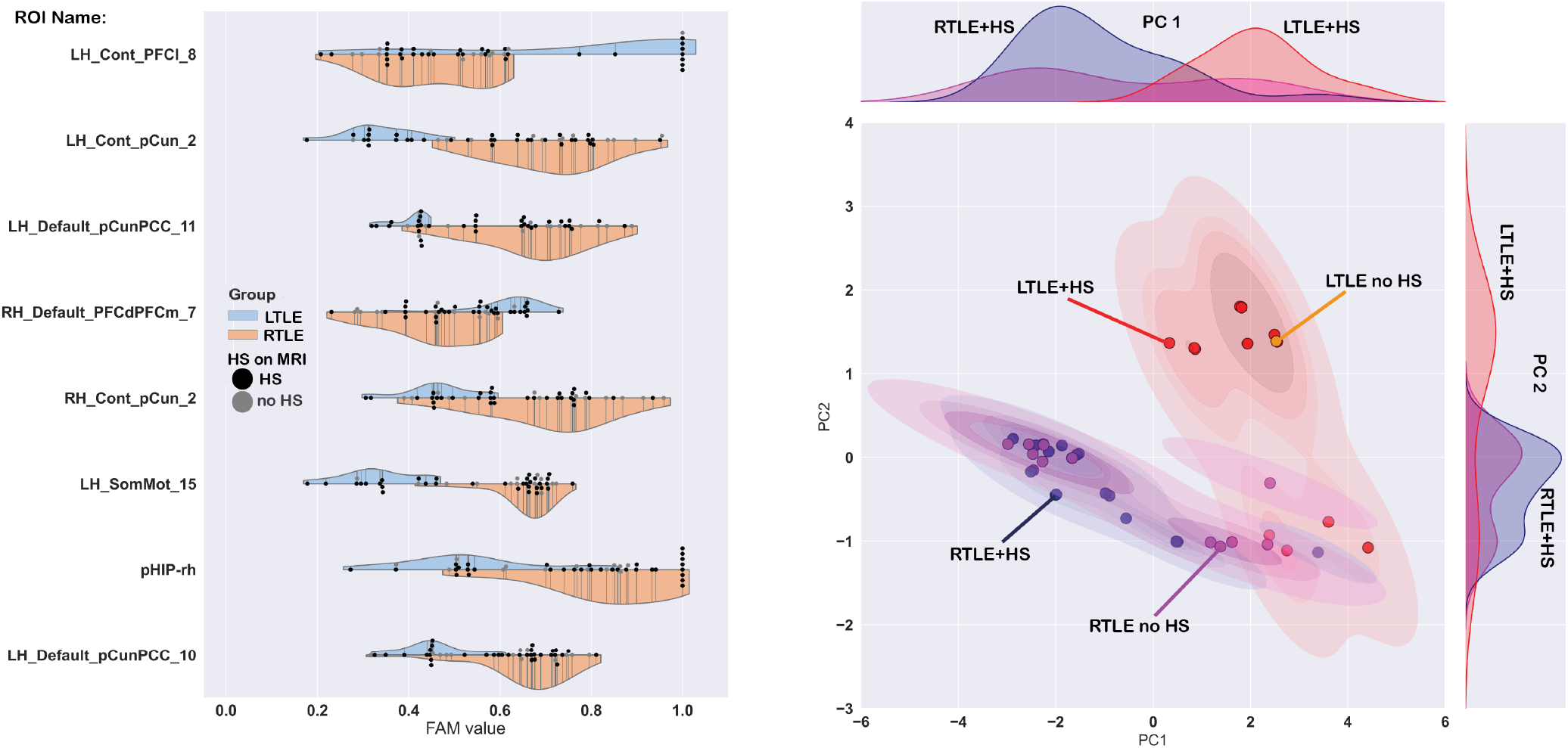
Group level anomalies in mesial temporal lobe epilepsy. **A)** Distribution of individual functional anomaly maps (FAMs) for eight selected regions of interest (ROIs) from the 432 ROIs, ranging from 0 to 1 (highest anomaly). Each vertical line represents a patient within the left (blue) or right (orange) mesial temporal lobe epilepsy groups (LTLE and RTLE). Black dots represent patients with hippocampal sclerosis (HS) in contrast to gray non-HS patients. Illustrated regions were selected as they demonstrated the highest importance in the SVM classifier trained to predict laterality. **B)** Low-dimensional representation of individual FAM for LTLE and RTLE patients. Principal components (PC) 1 and 2, the histograms in the axes of the figure, represented 71% of the 432-ROI FAM variability. Kernel density estimation lines surrounding datapoints on scatter plot illustrate the underlying probability density function of patients in each group, and their area of overlap.

## Discussion

In this study, we mapped the extent of functional anomalies in contrast to a normative sample at the group level. We detected robust spatiotemporal aberrations in regions distant from the ipsilateral mesial temporal structures, particularly in limbic and somatosensory networks. Our classification model resulted in near 100% accuracy in lateralizing mTLE patients based on individual FAM, derived from resting state scans. This lateralization accuracy also applied to those mTLE patients without evidence of HS on their clinical MRIs. Consistent with known structural changes in mTLE pathology, we measured high degrees of anomaly in mesial temporal (hippocampus and amygdala) subregions using individual FAMs. However, individualized mesial temporal anomalies were less lateralizing compared to medial parietooccipital regions. While showing high accuracy for lateralization, we did not find an association between FAM patterns and surgical outcomes in our dataset.

### Functional aberrations beyond the seizure onset zone are revealed by FAM

Using both the group and individual FAM approaches, we identified a number of anomalous temporal and extra-temporal cortical regions in addition to the expected anomaly in mesial temporal regions. These results align with prior structural and functional studies supporting a network pathophysiology in mTLE: while ipsilateral mesial temporal structures are the presumed epileptogenic focus and the surgical target for treatment, electrophysiological and imaging data show compelling evidence of seizure generation and propagation onetworks.[6, 16, 36-38] For instance, interictal FDG-PET hypometabolism in temporal and extratemporal cortices ipsilateral to affected side is common, and informative in clinical lateralization of mTLE.[39] Large multicenter studies show cortical atrophy observed in bilateral temporal, medial parieto-occipital, sensorimotor, and perisylvian/opercular regions.[10, 40-42]

Based on an observed increase in ipsilateral-contralateral hippocampal FC with longer disease duration, Morgan et al. suggested that contralateral hippocampus play a significant role in generating seizures after several years of epilepsy.[43] Our previous study using rsfMRI and a machine learning approach revealed common bilateral and wide-ranging FC metric changes in focal epilepsy, including in a subset analysis in TLE.[12] We found a higher proportion of altered within-network connections in DMN (i.e. DMN-DMN connections) and somatosensory networks compared to other within- and between-network connections. Using FAM in this study, we found the highest spatiotemporal anomaly in multiple ROIs from somatosensory network, and bilateral medial parietooccipital regions with ROIs affiliated with default mode, visual, and frontoparietal control networks, and anatomically related to precuneus and posterior cingulate regions in group FAMs and cumulative individual FAMs. (figures 2 and 4). Overall, the results from our spatiotemporal analysis approach converge with results from different prior methods of comparing TLE patients to controls, which support our hypothesis that FAM detects physiologically meaningful differences between patients and controls.

### Accurate lateralization of the seizure onset hemisphere using individual FAM

Our classification model showed very high accuracies in determining the laterality of temporal lobe of seizure onset. Although developed on a single-center dataset, we used a stratified splitting of the training and testing patients, and reported accuracy on participants unseen by the model across 1000 repetitions. While plotting in the reduced dimensions of the first two principal components illustrates the separation of individual FAMs based on their laterality, our use of a linear SVM model provides the opportunity to look at individual ROIs with highest importance in accurate classification (Figure 4). Remarkably, presence of HS does not appear to significantly contribute to lateralization accuracy. Lateralization is a crucial juncture in the path to surgery with the aim to cure epilepsy, and a common indication for invasive EEG recording. Although the presence of unilateral HS on clinical MRI improves the chance of seizure-freedom, around 20-30% of mTLE patients show seizure recurrence after selective amygdalohippocampectomy or anterior temporal lobectomy.[44] Discrepancies in outcome may in part point to a limitation in characterizing those with recent onset of disease or certain mTLE subtypes using anatomical MRI, both significant factors in planning treatment and predicting clinical outcomes. Overall, validating our non-invasive method of lateralization from rsfMRI data will complement clinical evaluation for mTLE patients with and without HS alike, and may lead to less need for invasive recording and better outcomes. Both group FAM and the combined individual FAM heatmap results highlight functional differences in RTLE and LTLE, when contrasted to the same control group, adding to evidence from previous structural and functional imaging studies that left- and right-onset focal epilepsies are not mere mirror images.[11, 12, 45] One possible explanation for right-left differences may relate to adaptive and compensatory changes in asymmetric networks such as language or even somatosensory networks.[46, 47] Functional connectivity studies support the hypothesis that presence of seizure focus in left hemisphere is associated with reorganization in language[48] and sensorimotor[49] networks as a developmental and age-related aspect of epilepsy. Overall, our findings highlight the impor-tance of avoiding mixing left- and right-hemispheric onset epilepsies in group analyses.

### Functional anomalies to complement structural changes

Structural changes, connectivity alterations, and functional changes in mTLE are related, but not completely “coupled.”[50, 51] Our results showed high FAM in subcortical, particularly mesial temporal regions, but a lower importance in lateralization model compared to many distant cortical regions in the DMN and somatosensory networks. Importantly, the lateralization models appear to perform similarly in patients with HS and without HS, a strong structural lateralizing finding. Our results suggest that rsfMRI analysis may provide an added value to current presurgical evaluation methods. Prior studies have studied rsfMRI in mTLE. One group reported an AUC of up to 0.87 with a regression model using connections between mesial temporal structures and DMN regions, and reported inferior parietal lobules and precuneus showing decreased connectivity in RTLE compared to LTLE patients.[19] Another study found a decrease in the functional thalamocortical connectivity in multiple posterior and ventromedial thalamic segments in both hemispheres.[18] Morgan et al. identified FC of the ventral lateral nucleus of the right thalamus to the bilateral hippocampi to distinguish left from right TLE patients.[52] Using ALFF, a direct measure of rsfMRI signal power, Sainberg et al. found a relative increase in bilateral anterior hippocampi, more prominent on the ipsilateral hemisphere for both LTLE and RTLE, which may correlate with disease severity and progression in mTLE. They also reported a lower ALFF in DMN regions in RTLE, but not LTLE patients.[21] Hwang et al. reported using ALFF for lateralization of TLE, reaching Leave-one-out cross-validation accuracies of 83% in training with specific frequency bands.[22] A recent study used a three-dimensional convolutional neural network trained on synthetically altered data for lateralization of mTLE, and reported a 90.6% accuracy in 32 patients. Most useful regions in classification based on a gradient-weighted class activation map included default mode, medial temporal, and dorsal attention networks.[53] In comparison, our method in this study uses a different machine learning approach to develop FAM, and a simple supervised classifier to achieved near 100% lateralization accuracy. Although we did not use traditional FC metrics and focused on measuring spatiotemporal alterations, this FAM approach maintains a connectivity context through extracting timeseries from functional cortical[28] and subcortical[29] ROIs. Based on our previous study using FC[12] and current results using FAM, it appears that cortical regions such as the heteromodal medial parieto-occipital as well as the unimodal somatomotor regions contribute to more accurate lateralization compared to subcortical structures, particularly the ipsilateral mesial temporal and limbic cortices. We may attribute this observation to the bilateral network dysfunction in mTLE and also high resting state variability in the healthy population. Bilateral amygdala and hippocampi are highly connected, and electrophysiological evidence suggests rapid spreading of epileptic activity to the contralateral side.[54, 55] While FAM may measure the degree of contralateral interictal dysfunction, compensatory engagement of the healthier temporal lobe[20] may contribute to measured aberrations as well. Larger and multimodal evaluation of hippocampal connections and functional anomalies may help test the above hypothesis.

### Technical advantages of FAM

In generating individual maps compared to a normative sample, FAM may provide different and unique information about individual patients compared to other methods. There are technical factors that make our FAM method suitable for clinical studies. First, we used an ROI- level FAM method, which in comparison to the original voxel-based method provides several computational advantages and makes transition to a clinical imaging tool more feasible. Instead of tens of thousands of time-series in voxel-based analysis, we used only 432 time-series in our models. Second, we used functionally-defined subcortical and cortical ROIs for averaging fMRI signal and contrasting patients’ to healthy individuals’ time-series. This physiologically meaningful “lumping” of data is also likely to improve signal to noise ratio, and add network-level context to the studied spatiotemporal properties. Our subcortical atlas provides additional granularity to analyses of functionally heterogenous regions relevant to mTLE. In particular, we utilize two segments for each hippocampus and amygdala, and four segments for each thalamus. Last, our method allows for future studies to choose custom segmentation schemes designed to test specific hypotheses. Our remarkable accuracy for lateralization in this dataset can be tested in focal non-TLE as well, as there is evidence for shared functional anomalies across all focal epilepsies. Although our dataset was not powered for detecting an association of FAM signal with surgical outcomes in this study, future studies with multimodal imaging data, pathology, and detailed clinical characteristics data may identify an association between individualize FAM and other clinical characteristics beyond laterality in mTLE. Larger datasets and prospective studies are required for validation of our findings before they can be considered as clinical tools.

## Limitations

In addition to the retrospective nature of the current analysis, there are other limitations. Our sample size is relatively small for most classification models. Some of the anomalies detected in TLE might be attributed to unmatched factors in controls including medication effects and comorbidities, however it is unlikely that are lateralizing. The validity of comparing FAM from different centers needs additional studies. In summary, we demonstrated spatiotemporal aberrations in TLE using a new ROI-based FAM method, and achieved a high accuracy lateralizing mTLE using individual FAM results. Future studies should use validation datasets, correlate FAM patterns with clinical course and symptoms, and treatment outcomes.

## Data Availability

Anonymized data and code will be shared upon request by qualified investigators.

## ACKNOWLEDGEMENTS

This study was supported by the National Institutes of Health (NIH) National Center for Advancing Translational Sciences (UL1TR001876/KL2TR001877 to T.G.), NIH National Institute of Neurological Disorders and Stroke (NS075270, NS110130, NS108445 to V.L.M., NS097618 and NS112252 to D.J.E) and the NIH National Institute of Child Health and Human Development (Intellectual and Developmental Disabilities Research Center 1U54HD090257-01). None of the authors has any conflict of interest to disclose.

